# Matrix metalloproteinase 2 and 9 enzymatic activities are selectively increased in the myocardium of Chronic Chagas Disease cardiomyopathy patients: role of TIMPs

**DOI:** 10.1101/2021.12.17.21267825

**Authors:** Monique Andrade Baron, Ludmila Rodrigues Pinto Ferreira, Priscila Camillo Teixeira, Ana Iochabel Soares Moretti, Ronaldo Honorato Barros Santos, Amanda Farage Frade, Andréia Kuramoto, Victor Debbas, Luiz Alberto Benvenuti, Fabio Antônio Gaiotto, Fernando Bacal, Pablo Pomerantzeff, Christophe Chevillard, Jorge Kalil, Edecio Cunha-Neto

**Affiliations:** Laboratory of Immunology, Heart Institute (InCor), University of São Paulo, School of Medicine, 05403-900, São Paulo, Brazil; Division of Clinical Immunology and Allergy, University of São Paulo, School of Medicine, 01246100, São Paulo, Brazil; Institute for Investigation in Immunology (iii), INCT, 05403-001, São Paulo, Brazil; Universidade Santo Amaro (UNISA), São Paulo, Brazil.; Vascular Biology Laboratory, Heart Institute (InCor), University of São Paulo, School of Medicine, 05403-900, São Paulo, Brazil; Division of Transplantation, Heart Institute (InCor), University of São Paulo, School of Medicine, 05403-900, São Paulo, Brazil; INSERM, UMR_1090, Aix Marseille Université, TAGC Theories and Approaches of Genomic Complexity, Institut MarMaRa, Marseille, France

**Keywords:** Chagas disease, metalloproteinases, heart failure, cardiac remodeling, MMPs

## Abstract

Chronic Chagas disease (CCC) is an inflammatory dilated cardiomyopathy with a worse prognosis compared to other cardiomyopathies. We show the expression and activity of Matrix Metalloproteinases (MMP) and of their inhibitors TIMP (tissue inhibitor of metalloproteinases) in myocardial samples of end stage CCC, idiopathic dilated cardiomyopathy (DCM) patients, and from organ donors. Our results showed significantly increased mRNA expression of several MMPs, several TIMPs and EMMPRIN in CCC and DCM samples. MMP-2 and TIMP-2 protein levels were significantly elevated in both sample groups, while MMP-9 protein level was exclusively increased in CCC. MMPs 2 and 9 activities were also exclusively increased in CCC. Results suggest that the balance between proteins that inhibit the MMP-2 and 9 is shifted toward their activation. Inflammation-induced increases in MMP-2 and 9 activity and expression associated with imbalanced TIMP regulation could be related to a more extensive heart remodeling and poorer prognosis in CCC patients.

## Introduction

Chronic Chagas cardiomyopathy (CCC) is the most important chronic clinical manifestation of Chagas disease, a zoonosis caused by the protozoan parasite, *Trypanosoma cruzi* (*T. cruzi*) naturally transmitted through haematophagous reduviid bugs, but also by blood transfusions, organ transplants or congenitally [1]. The World Health Organization estimates that approximately 7-8 million people are infected with *T. cruzi* in Central and South America [2], with an estimated 300.000 cases in the United States and 120.000 cases in Europe alone due to migration of people from endemic areas. The acute phase of Chagas disease usually lasts 6-8 weeks, and most frequently is oligo- or asymptomatic [3]. About 30% of the asymptomatic patients will develop CCC, a progressive, fibrotic disease in which myocardial inflammation plays a fundamental role causing tissue damage and also dysregulation of heart extracellular matrix (ECM) leading to a maladaptive cardiac remodeling that constitutes the basic step to a progressive heart failure [4–8].

The term cardiac remodeling describes a response mechanism in response to injury and differs from the physiological cardiac hypertrophy observed during gestation and intensive exercising [9, 10]. Different cardiovascular disorders, like myocardial infarction, ischemia, reperfusion injury and viral myocarditis displays an intense cardiac remodeling that involves cardiac fibroblasts, the main cell type responsible for ECM regulation and homeostasis [6, 11]. Cardiac fibroblasts perform important functions by synthetizing ECM molecules as well as regulating the ECM turnover by secreting matrix metalloproteinases (MMPs) [6, 12]. MMPs are zinc-dependent endopeptidases responsible for degradation and remodeling of ECM components including collagens, proteoglycans, fibronectin and laminin within the injury zone [12]. Because MMPs have a dual role functioning in both physiological and pathological processes, their functions should be tightly regulated. MMPs are synthesized and secreted, in most cases, as inactive proenzymes (pro-MMPs) that are activated by other proteinases [13, 14]. MMPs also regulate the activities of effector proteins involved in inflammation and fibrosis [15]. The homeostasis in a normal healthy heart is dependent on the balance between activation of pro-MMPs to active degradative MMPs, a process controlled by complex formation with their endogenous inhibitors: tissue inhibitors of MMPs (TIMPs) [13, 16]. MMPs expression and activity were shown to be dysregulated in different cardiovascular diseases and in failing hearts of various etiologies like acute myocardial infarction, DCM and heart failure [17]. Two important members of MMPs, MMP-2 and MMP-9, are from the group of gelatinases and have been indicated as key enzymes responsible for profound cardiac remodeling [18, 19]. Right after an acute injury, like a myocardial infarction, the pre-existing pro-MMP-2 and 9 within the myocardium are activated and disrupt the fibrillar collagen network to allow inflammatory cells to infiltrate into the infarct zone to remove the necrotic cells [20]. Myocytes, fibroblasts and activated macrophages as well as neutrophils are thought to be the main cellular sources for the increased MMP-2 and MMP-9 levels [21]. Previous studies have shown higher serum and plasma levels of MMP-2 and MMP-9 in CCC [22, 23]. Other MMPs, such as MMP-3, MMP-8, MMP-13 and MMP-12 also have as substrate collagen type I and III, main components of heart extracellular matrix [24, 25]; Extracellular matrix metalloproteinase Inducer EMMPRIN, an inducer of MMP2 and MMP9 [26, 27]. Here, we show for the first time, gene and protein expression, as well as enzymatic activity of MMP-2, MMP-9 and gene and protein expression of their specific inhibitors TIMP 1, 2, 3 and 4 and EMMPRIN in myocardial tissue samples from CCC patients compared to DCM and from heart transplant donors (control samples). Our results suggest an important role of MMP-2 and MMP-9 in myocardial remodeling and in cardiac dysfunction observed in patients with CCC.

## Materials and Methods

### Ethics Statement

The protocol was approved by the Institutional Review Board of the School of Medicine, University of São Paulo (Protocol number 492/682) and written informed consent was obtained from the patients. In the case of samples from heart donors, written informed consent was obtained from their families.

### Samples of human myocardium

Myocardial samples were obtained from left ventricular-free wall heart tissue from end-stage heart failure patients at the moment of heart transplantation. The patients belonged to three diagnostic groups: CCC (at least 2 positive results in 3 independent anti-T. cruzi serology tests – ELISA immunoassay, indirect immunofluorescence assay and indirect hemagglutination test), DCM (idiopathic dilated cardiomyopathy) and controls, left ventricular free wall samples obtained from healthy hearts of organ donors, which were not used for transplantation due to size mismatch with available recipients (**Table 1**). All Chagas disease patients underwent standard electrocardiography and echocardiography. Echocardiography was performed in the hospital setting using an Acuson Sequoia model 512 echocardiographer (Siemens, Germany) with a broad-band transducer. The left ventricular dimensions and regional and global function evaluations were performed using a 2 dimensions and M mode approach, in accordance with the recommendations of the American Society of Echocardiography. Patients with CCC presented with abnormal electrocardiography findings that ranged from typical conduction abnormalities (right bundle branch block and/or left anterior division hemiblock) to severe arrhythmia [28, 29]. A group of patients also presented varying degrees of ventricular dysfunction classified on the basis of left ventricular ejection fraction with all other causes of ventricular dysfunction/heart failure excluded. Characteristics of patients and normal donors whose samples were used in this study are described in **Table 1** and **Supplemental Table 2**.

**Table 1.** Clinical characteristics of the study subjects.

| CLINICAL PARAMETERS | GROUPS |  |  |
| --- | --- | --- | --- |
|  | CCC | DCM | CONTROL |
| Age | 47.8 ±14,2 | 40,6±17,1 | 28,3 ± 11,0 |
| Male, n (%) | 6 (35%) | 9 (75%) | 6 (100%) |
| Sample size, n | 17 | 12 | 6 |
| Body Surface (m<sup>2</sup>) | 1.6 ± 0.3 | 1.7 ± 0.1 | N.D. |
| Ejection Fraction- EF (%) | 23.59 ± 8.5 | 21.6 ± 7.9 | N.D. |
| Fractional shortening (%) | 11.94 ± 7.3 | 12.2 ± 3.7 | N.D. |
| Left ventricular end diastolic diameter LVEDD (mm) | 62.12 ± 98 | 70.6 ± 19.8 | N.D. |
| Left ventricular end systolic diameter LVESD (mm) | 48.82 ± 15.6 | 62.7 ± 20.1 | N.D. |
| Ventricular Septum (mm) | 8.53 ± 1.7 | 8.3 ± 1.4 | N.D. |
| Posterior wall of left ventricle (mm) | 8.5 ± 1.8 | 8.6 ± 1.4 | N.D. |
| Left atrial diameter (mm) | 49.18 ± 5.5 | 52.1 ± 7.4 | N.D. |
| Aortic sinus (mm) | 29.53 ± 3.1 | 31.8 ± 5.3 | N.D. |
| Myocardial mass index (g/m<sup>2</sup>) | 187.88 ± 69.3 | 233.8 ± 76.6 | N.D. |
| Relative wall thickness (mm) | 0.68 ± 0.5 | 0.60 ± 0.1 | N.D. |
| Diastolic volume (ml) | 288.41 ± 136.6 | 349.9 ± 131.8 | N.D. |
| Systolic volume (ml) | 194.06 ± 131.8 | 267.5 ± 127.4 | N.D. |

**Table 1.** Clinical characteristics of the study subjects.
| CLINICAL PARAMETERS | GROUPS |  |  |
| --- | --- | --- | --- |
|  | CCC | DCM | CONTROL |
| Age | 47.8 ±14,2 | 40,6±17,1 | 28,3 ± 11,0 |
| Male, n (%) | 6 (35%) | 9 (75%) | 6 (100%) |
| Sample size, n | 17 | 12 | 6 |
| Body Surface (m<sup>2</sup>) | 1.6 ± 0.3 | 1.7 ± 0.1 | N.D. |
| Ejection Fraction- EF (%) | 23.59 ± 8.5 | 21.6 ± 7.9 | N.D. |
| Fractional shortening (%) | 11.94 ± 7.3 | 12.2 ± 3.7 | N.D. |
| Left ventricular end diastolic diameter LVEDD (mm) | 62.12 ± 98 | 70.6 ± 19.8 | N.D. |
| Left ventricular end systolic diameter LVESD (mm) | 48.82 ± 15.6 | 62.7 ± 20.1 | N.D. |
| Ventricular Septum (mm) | 8.53 ± 1.7 | 8.3 ± 1.4 | N.D. |
| Posterior wall of left ventricle (mm) | 8.5 ± 1.8 | 8.6 ± 1.4 | N.D. |
| Left atrial diameter (mm) | 49.18 ± 5.5 | 52.1 ± 7.4 | N.D. |
| Aortic sinus (mm) | 29.53 ± 3.1 | 31.8 ± 5.3 | N.D. |
| Myocardial mass index (g/m<sup>2</sup>) | 187.88 ± 69.3 | 233.8 ± 76.6 | N.D. |
| Relative wall thickness (mm) | $0.68 \pm 0.5$ | $0.60 \pm 0.1$ | N.D. |
| Diastolic volume (ml) | $288.41 \pm 136.6$ | $349.9 \pm 131.8$ | N.D. |
| Systolic volume (ml) | $194.06 \pm 131.8$ | $267.5 \pm 127.4$ | N.D. |

N.D.: not done.
Control: Heart donors; DCM: Idiopathic dilated cardiomyopathy; CCC: Chronic Chagas cardiomyopathy.
Datas were expressed as mean ± SD.
Reference values: EF: Ejection Fraction ( $>55\%$ ); Ejection Fraction - EF (25-43%); Left ventricular end diastolic diameter - LVESD (mm): (42-59mm); Left ventricular end systolic diameter – LVESD (mm): (25-39mm); Ventricular Septum (mm): (6-10mm); Posterior wall of the left ventricle (mm): (6-10mm); Left atrial diameter (mm): (30-40mm); Aortic sinus (mm): (31-47mm); Myocardial mass index ( $\text{g}/\text{m}^2$ ): (49-115 $\text{g}/\text{m}^2$ ); Relative wall thickness (mm): ( $<0.42$ ).

### Samples preparation

All samples were cleared from pericardium and fat and 100mg pieces of left ventricle heart wall were quickly frozen in liquid nitrogen and stored at −80°C and another set was fixed in a solution of 10% neutral buffered formalin, embedded in paraffin and sectioned at thickness of individual 5 µm sections. The slides were stained with Hematoxylin-Eosin (H&E) for evaluation of the intensity and location of the inflammatory infiltrate and PicroSirius Red (Abcam, UK) staining to visualize the degree of fibrosis. Stained sections were acquired with a 20x objective lens by Leica DM2500 microscope (Leica Cambridge, Ltd, UK) and true color digital images were captured. Slides were scored for the intensity of myocarditis and for the presence or absence of hypertrophy (**Supplemental Table 2**).

### Morphometric analysis for myocardial collagen area

A total of 15 images were captured per myocardium tissue sample under 10X magnification; we studied 5 samples of each patient group (CCC, DCM, and control). For quantitative evaluation of fibrosis, collagen area was calculated using Leica Qwin Plus 3.5 image analysis software, as percentage of fibrosis area per total area of heart tissue sections.

### Isolation of total RNA

Total RNA was isolated from 100mg of tissue from each heart sample (CCC N=17, DCM N=12, Control N=6) by mechanical disruption with the Precellys 24-bead-based homogenizer (Bertin Technologies, France) using 3 cycles of 15 seconds with pause of 20 seconds each at 6.000 rpm. Samples were homogenized in 1ml of Trizol reagent (Life Technologies, USA) following the manufacturer’s protocol. All sample had an OD 260/280 ration ≥1.9. The RNA integrity was analyzed on a Bioanalyzer 2100 (Agilent, USA). Only samples with RIN (RNA Integrity Number) value > 6.5 were used in our analysis.

### Analysis of mRNA expression by quantitative real time polymerase chain reaction (Real Time – qPCR)

RNA from each sample was treated with Rnase-free DNAse I (United States Biological, Ohio, USA) cDNA was obtained from 5 µg total RNA using Super-script II reverse transcriptase (Invitrogen, USA) and mRNA expression was analyzed by real-time qPCR with SYBR Green I PCR Master Mix (Applied Biosystems, USA) with 250nM of sense and anti-sense specific primers using the ABI Prism 7500 Real Time PCR System. The following primers were designed using Primer Express software version 3.0 (Applied Biosystems, USA; **Supplemental table 1**). All the samples were processed in triplicate for the target and for the endogenous reference gene, RPLP0. A set of “no RT” and "non template controls" controls were included in each experiment. Ct values were averaged for the replicates and expression was calculated as the mean ± interquartile ranges per group for each individual data point using the relative expression equation (fold change over control samples) by the 2^-DDCT^ method.

### Protein extraction from heart tissue samples

Protein was isolated from each heart sample with RIPA buffer containing 150 mM sodium chloride, 1% sodium deoxycholate, 0.1% sodium dodecyl sulfate, 100 mMl/Tris-HCL pH 7.5, and proteolytic enzyme inhibitors (1mM phenymethylsulfonylfluoride, PMSF and 1% aprotinin) (Sigma-Aldrich, USA) by mechanical disruption with the Precellys 24-bead-based homogenizer using 3 cycles of 15 seconds with pause of 20 seconds each at 6,000 rpm at 4°C. The homogenate was sonicated for 3 cycles of 10 seconds each of 10 Watts (60 Dismembrator Sonic, Fischer Scientific, USA), centrifuged at 12.000g for 30 minutes at 4°C. Supernatants were collected and protein quantification was performed using the Bradford method (Bio-Rad Laboratories, USA).

### Protein expression analysis by Western blotting

About 70µg of isolated protein from each myocardial sample (5 samples per group: CCC, DCM and Control) were heated for 5 minutes at 95°C, and subjected to one dimensional electrophoresis (SDS-PAGE) using 12.5% polyacrylamide using the vertical electrophoresis System Ruby SE600 (GE Healthcare, USA). After electrophoresis, proteins were transferred from gel to a nitrocellulose membrane using the TE Semi-Dry Transfer Unit (GE Healthcare, USA). The nitrocellulose membranes (Bio-Rad, Laboratories, USA) were blocked for nonspecific binding with 5% non-fat dry milk in Tris-buffered saline and 0.1% Tween 20% (TBST) for 2h at room temperature. Blots were washed in TBST twice over 30 minutes and the ECL Plus Western Blotting Detection Reagents, were used for detection. Images were capture using the ImageQuant LAS 4000 equipment (GE Healthcare, USA) and membranes were imaged using a LI-Cor Odyssey scanner (LI-COR Biotechnology - UK Ltd). Analysis of densitometry was performed using the program ImageQuant TL, and the data was normalized to beta-actin.

### Zymography assay analysis of MMP-2 and MMP-9 activity

Activities of MMP-2 and MMP-9 in myocardium tissue sample from CCC, DCM and Control (5 samples/each group) were examined by gelatin zymography. About 70µg protein isolated from each heart tissue sample were applied to 1D electrophoresis using 10% polyacrylamide gel copolymerized with 1% gelatin type A from porcine skin (Sigma, USA) as a substrate for gelatinolytic proteases. After electrophoresis, the gels were washed twice in 2.5% Triton X-100 (Amersham Biosciences, UK) in water for 30 minutes. The gels were then incubated overnight at 37°C in reaction buffer (5mM CaCl_2_, 2mM NaN_3_ and 50mM Tris-HCL buffer pH 7.4). After incubation, the gels were stained for 3h in 45% methanol,10% glacial acetic acid containing 1% Coomassie Blue R-250 (Amersham Biosciences/GE Healthcare, USA) and were subsequently partially destained with the same solution without dye. The gelatinolytic activity of each MMP was qualitatively evaluated as a clear band of digested gelatin against a blue background. To confirm that the visualized bands of lysis were due to MMP activity, selected duplicate gels was incubated with MMP inhibitors (5mM EDTA or 10mM 1,10-phenanthroline). Proteolytic signals of active MMP-2 and MMP-9 were quantified based on the peak area, using the software ImageQuant TL [30].

### Statistical analysis

Difference between groups was determined by one-way Analysis of Variance (ANOVA) followed by a non-parametrical test (Mann-Whitney Rank Sum Test). All statistical analyses were performed with GraphPad Prism 6.0 software (GraphPad Prism Software, Inc., USA). Results were expressed as mean ± SEM. P*-*values were considered significant if ≤ 0.05 (marked with a * symbol).

## Results

### Cardiomyocyte hypertrophy and collagen deposit in heart of CCC and DCM patients

The histopathological analysis revealed that both groups CCC and DCM present cardiomyocyte hypertrophy with nuclear enlargement (**Figure 1A**). As expected, myocarditis associated with a predominant lymphocytic infiltration was observed only in heart samples of CCC patients, evidenced by H&E staining (**Supplemental Table 2, Figure 1A**). To evaluate ECM fibrosis distribution, heart tissue sections were stained with Picro Sirius Red to detect collagen deposition. Both CCC and DCM displayed a significantly higher percentage of fibrosis area as compared to Controls (**Figure 1B**). Collagen area was also significantly higher in the heart tissue sections from CCC compared to DCM group. There were no significant differences in the analyzed clinical parameters such as ejection fraction (E.F), left ventricle diastolic diameter (LVDD) and left ventricle systolic diameter (LVSD) between DCM and CCC groups (**Table 1**).

**Figure 1.**
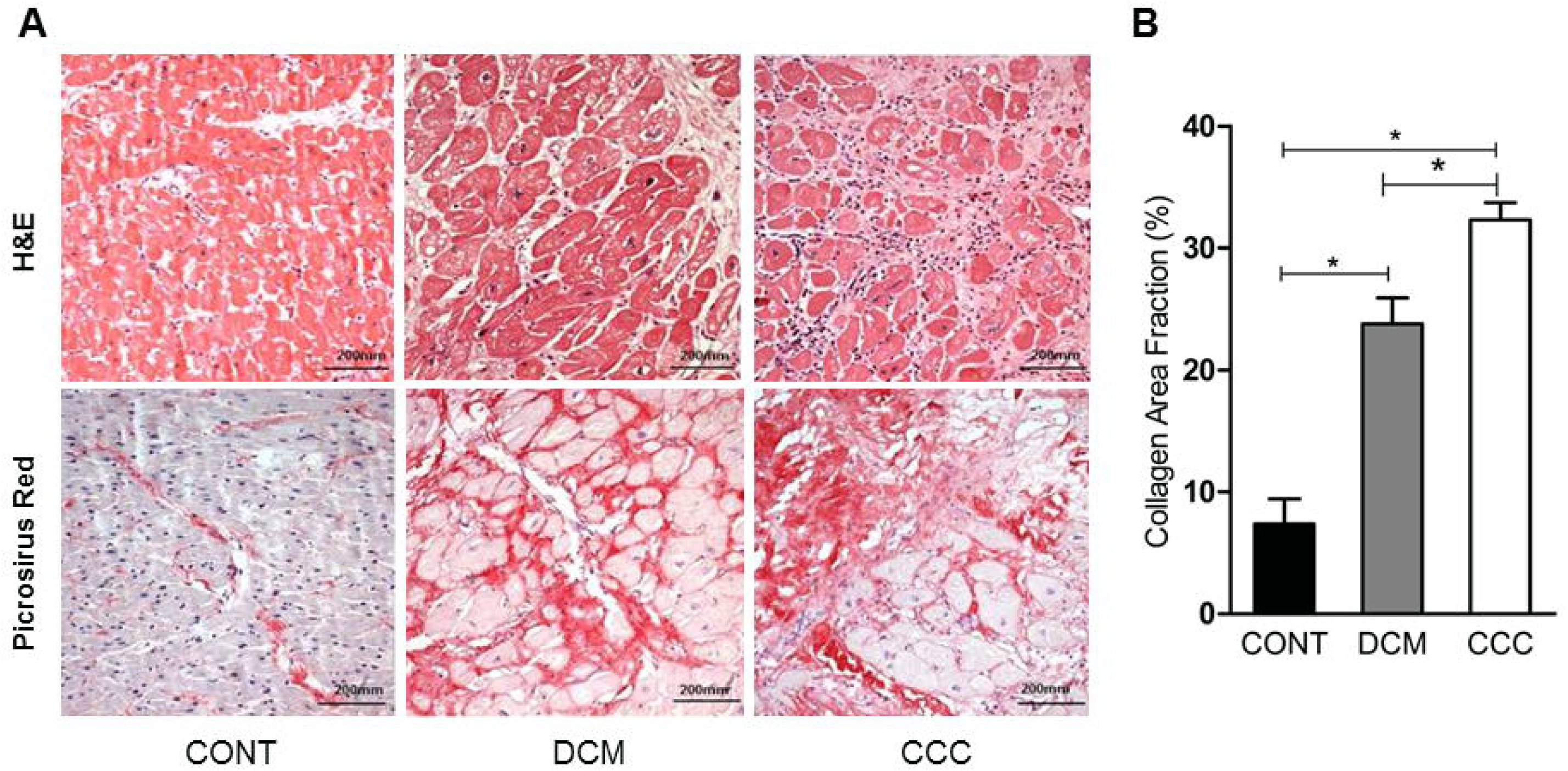
Histopathological features of myocardial samples. **(A)** Slides of hematoxylin-eosin (H&E) and picrosirius red stained myocardial sections of representative patients with CCC, DCM and individuals without cardiomyopathies (Controls). Representative heart sections were shown with magnification: 20X. Myocardial hypertrophy characterized by fiber and nuclear enlargement is evident in the CCC and DCM groups. Lymphocytic myocarditis is present only in the CCC group. Interstitial fibrosis stained in red with picrosirius red is present in the CCC and DCM groups. **(B)** The severity of fibrosis was quantified by collagen area fraction (%) in DCM and CCC group exhibit a significant increase in the deposition of collagen. Groups were compared by a non-parametrical test (Mann-Whitney Rank Sum Test) with GraphPad Prism software (version 6.0; GraphPad). Results were expressed as mean±SEM. *\* P*-values were considered significant if p value (0.05) corrected for multiple comparisons by Bonferroni’s method (corrected p value=0.05/3=0.0166).

### MMPs are significantly altered in heart from CCC and DCM patients

We evaluated gene and protein expression of different classes of MMPs (MMP-2, MMP-3, MMP-8, MMP-9, MMP-12, MMP-13) and EMMPRIN in heart samples from DCM, CCC and CONT patients. We observed a significant upregulation in gene expression of MMP-2, MMP-9 (**Figure 2A**), MMP-12, MMP-13 and EMMPRIN in both DCM and CCC heart samples compared to Control (**Supplemental figure 1**). MMP-3 and MMP-8 mRNA expression were undetectable in all samples tested. Western blotting analysis showed that MMP-2 protein is increased in both DCM and CCC heart samples and MMP-9 protein is exclusively increased in CCC compared to Control group (**Figure 2B**). There were no significant differences in protein expression of MMP-3, MMP-8, MMP-12, MMP-13 and EMMPRIN between CCC, DCM and control samples (**Figure 6D**). Gelatin zymography was carried out on DCM, CCC and control patient heart samples (5 samples per group) to assess MMP-2 and MMP-9 enzymatic activity. **Figure 2C** shows the detection of gelatinolytic bands for MMP-2 (62kDa) and MMP-9 (88kDa) on SDS-PAGE gel. Densitometry analysis (**Figure 2D**) shows that both enzymes MMP-2 and MMP-9 are significantly more active in CCC heart tissue samples as compared to both control and/or DCM samples. MMP-2 and MMP-9 activities in the DCM group were not significantly different from control tissue.

**Figure 2.**
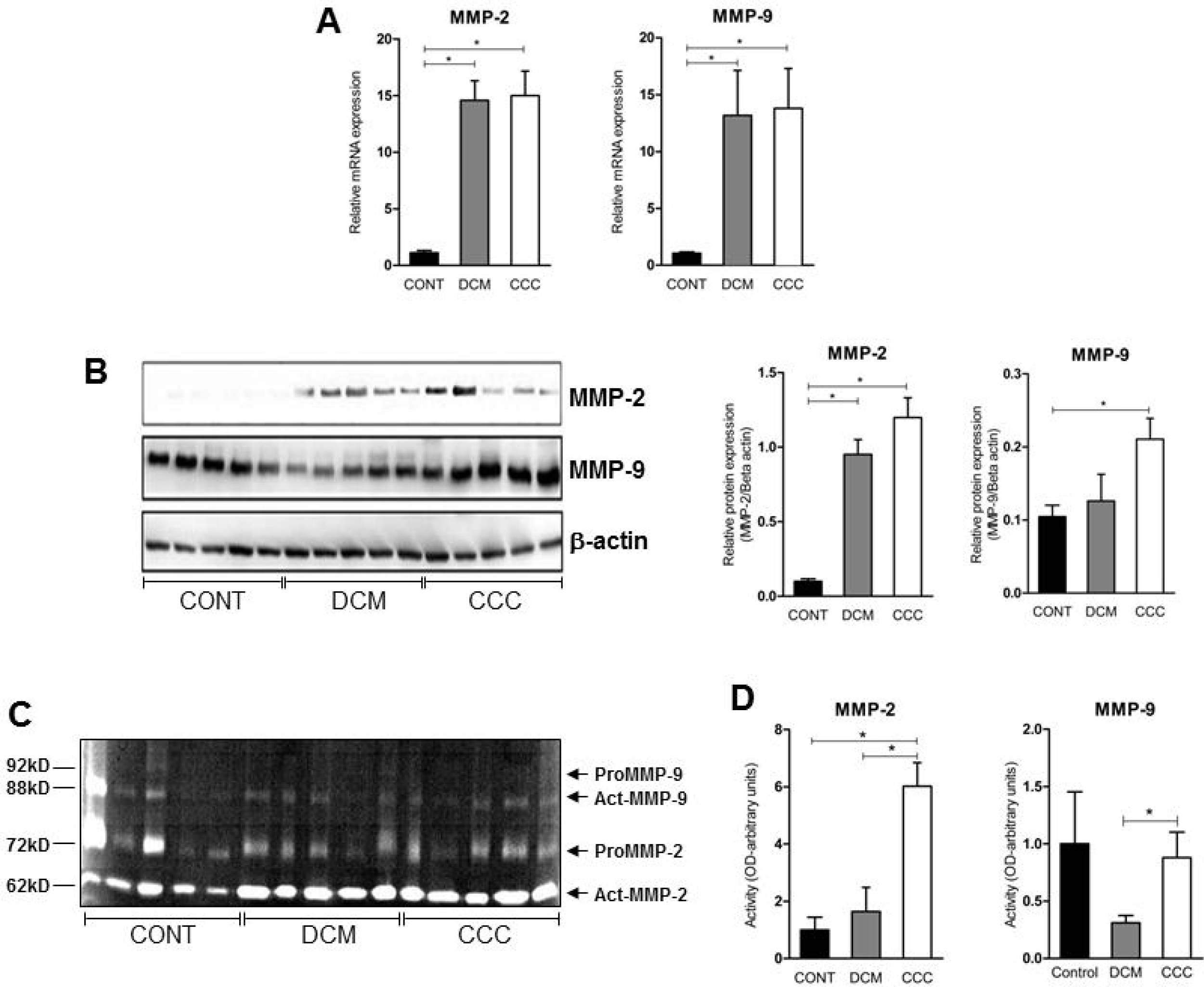
MMP-2 and MMP-9 mRNA and protein expression and activity in myocardium tissue from control, DCM and CCC samples. Myocardial expression of MMP-2 and MMP-9 mRNA. Real-time PCR analysis of mRNA expression in control (n=6), DCM (n=12) and CCC (n=17) myocardium. The expression was calculated as the mean ± SEM for each group as individual data points using the relative expression (fold change over control) by the 2^−ΔΔCt^ method, normalized with the endogenous gene RPL0, as described in Methods section. **(A)** Relative expression of MMP-2 mRNA and MMP-9 mRNA. **(B)** Western blotting image showing protein of MMP-2 and MMP-9 in extracts from myocardium samples from the control, DCM and CCC (n= 5 in each group). The densitometric values of each protein for each sample were normalized by the values of Beta actin, as described in methods section. Densitometry analysis of the MMP-2. Densitometry analysis of the MMP-9. **(C)** Zimography image showing activity of MMP-2 and MMP-9 in extracts from myocardium samples from control, DCM and CCC (n= 5 in each group). **(D)** Densitometry analysis of the activated MMP-2 (66 kD band) and activated MMP-9 (88kD band) results. Groups were compared by a non-parametrical test (Mann-Whitney Rank Sum Test) with GraphPad Prism software (version 6.0; GraphPad). Results were expressed as mean±SEM. *\* P*-values were considered significant if p value (0.05) corrected for multiple comparisons by Bonferroni’s method (corrected p value=0.05/3=0.0166). Primary antibodies (with their respective dilution) against the following proteins were used: MMP-2 (mouse monoclonal, 1:1000, Abcam, UK); MMP-3 (rabbit polyclonal, 1: 500, Abcam, UK); MMP-8 (rabbit polyclonal, 1:500, UK); MMP-9 (rabbit polyclonal, 1:1000, UK); MMP-12 (rabbit polyclonal, 1:1000, UK); MMP-13 (rabbit polyclonal, 1:1000, UK); EMMPRIN (mouse monoclonal, 1:500, Santa Cruz, Biotechnology, USA); TIMP-1 (1:1000); TIMP-2 (1:1000); TIMP-3 (goat plyclonal,1:500, Santa Cruz Biotechnology, USA); TIMP-4 (goat plyclonal,1:500, Santa Cruz Biotechnology, USA); Reck (mouse polyclonal,1:1000, Abcam, UK). Anti-beta-actin antibody (mouse monoclonal, 1:2000, Sigma, USA), was used to detect beta-actin, used as protein loading control. All antibodies were diluted in TBST and incubated at 4°C overnight. After washing twice over 30 minutes with TBST, each membrane was incubated with compatible secondary antibodies horseradish peroxidase conjugate (goat anti-rabbit, rabbit anti-goat or goat anti-mouse, 1:10000, Calbiochem, USA) for 2h at room temperature.

### Gene and Protein expression of Tissue inhibitors of MMPs (TIMPs) in heart from CCC and DCM patients

We assessed gene and protein expression of TIMPs 1, 2, 3 and 4 in heart tissue from CCC, DCM and control samples. A significant upregulation in gene expression was found for all TIMPs analyzed in CCC and DCM heart tissue samples (**Figure 3A**) compared to the control group (P≤ 0.05). TIMP-2 was the only inhibitor with significantly increased protein levels, both in DCM and CCC heart tissue samples as compared to Control samples (**Figure 3B**).

**Figure 3.**
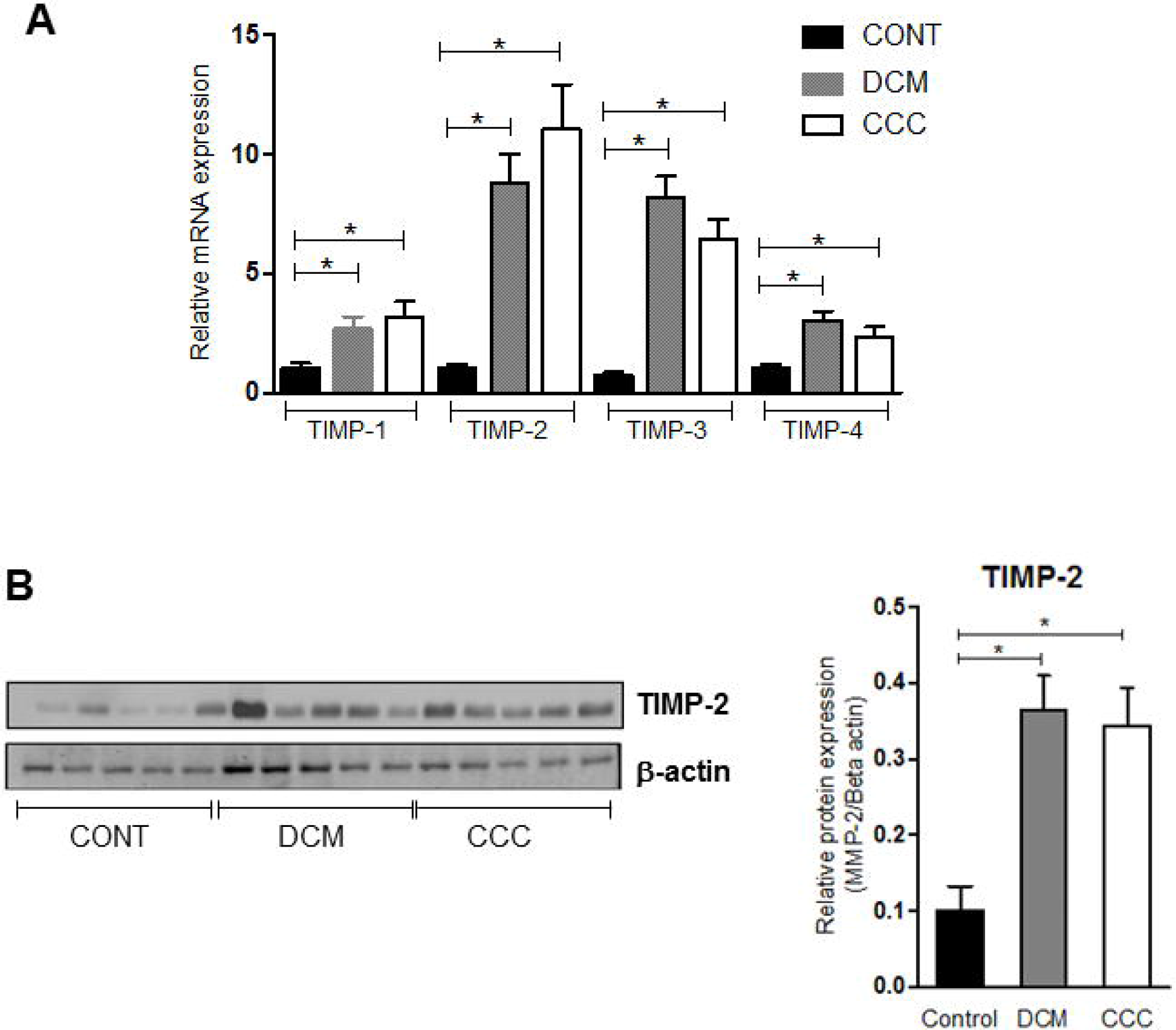
Tissue inhibitors of MMPs (TIMPs) mRNA expression and protein expression in heart tissue from CCC and DCM samples. Real-time PCR analysis of mRNA expression in control (n=6), DCM (n=12) and CCC (n=17) myocardium. The expression was calculated as the mean ± SEM for each group as individual data points using the relative expression (fold change over control) by the 2^−ΔΔCt^ method, normalized with the endogenous gene RPL0, as described in Methods section. **(A)** Relative expression of TIMP-1 mRNA; TIMP-2 mRNA; TIMP-3 mRNA; TIMP-4 mRNA. **(B)** Western blotting image showing protein of the 21 kDa TIMP-2 bands in extracts from myocardium samples from the control, DCM and CCC (n= 5 in each group). The densitometric values of TIMP-2 protein for each sample were normalized by the values of 42 kD Beta actin band, as described in methods section. Groups were compared by a non-parametrical test (Mann-Whitney Rank Sum Test) with *GraphPad Prism* software (version 6.0; GraphPad). Results were expressed as mean±SEM. *\* P*-values were considered significant if p value (0.05) corrected for multiple comparisons by Bonferroni’s method (corrected p value=0.05/3=0.0166).

### Balance between MMP-2 and MMP-9 and their tissue inhibitors (TIMPs) is shifted towards activation in CCC and DCM heart tissue

We observed that the MMP-2/TIMP-1, MMP-2/TIMP-2, MMP-2/TIMP-3 and MMP-2/TIMP-4 ratios were increased in CCC heart tissue compared to DCM or control group samples (P≤ 0.05) (**Figures 4A to 4D**). As shown in **Figures 5A** to **5C,** MMP-9/TIMP-1 and MMP-9/TIMP-3 ratios were also increased in CCC heart tissue in comparison to DCM or control group (P≤ 0.05); MMP9/TIMP2 and MMP-9/TIMP-4 ratio did not show any significant changes between the analyzed groups (**Figure 5D**).

**Figure 4.**
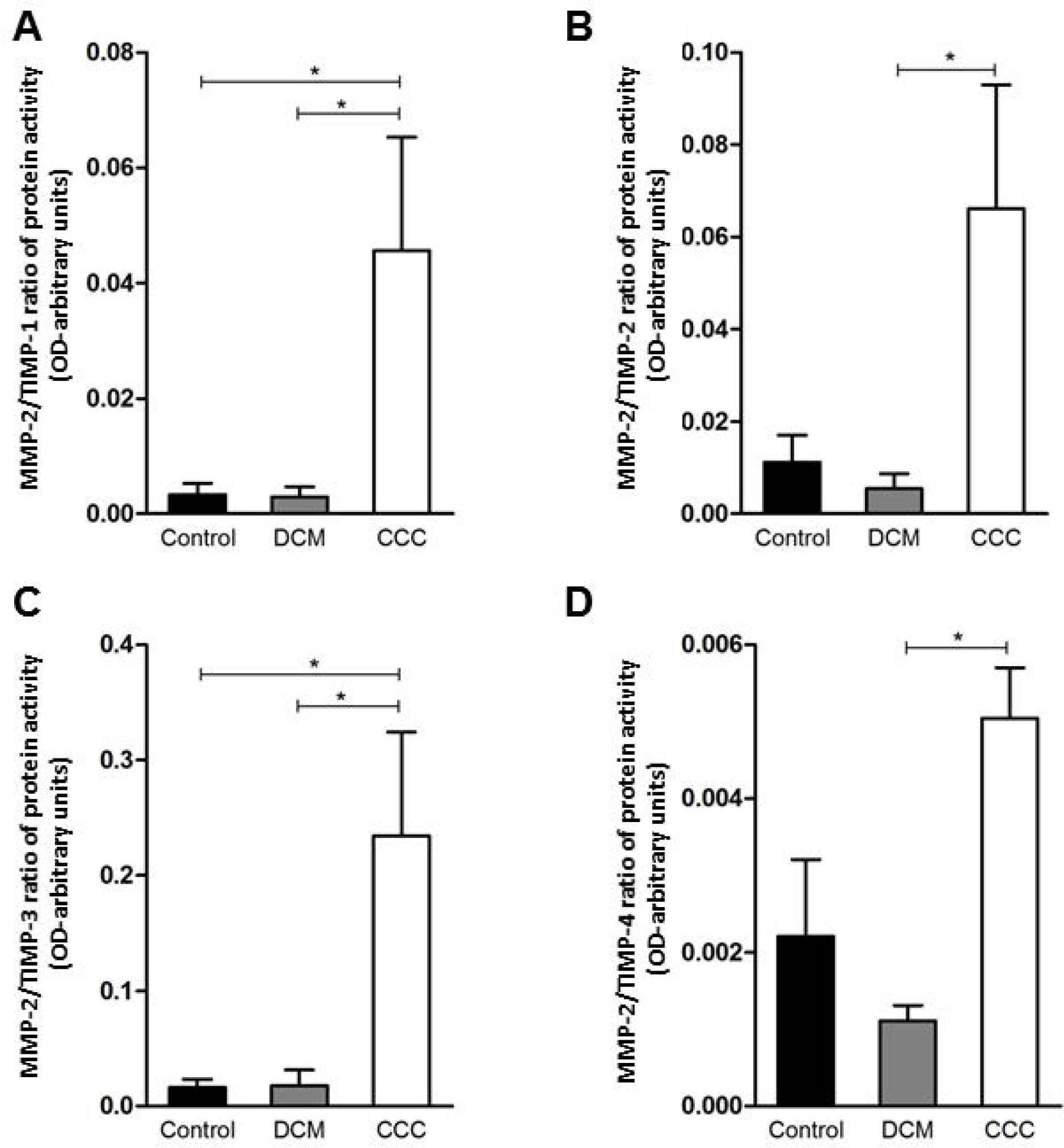
Ratios of MMP2/TIMP in heart tissue from CCC and DCM samples. Activity of MMP-2 / expression protein TIMP stoichiometric ratios were calculated for samples CCC, DCM and control. **(A)** MMP-2/TIMP-1. **(B)** MMP-2/TIMP-2. **(C)** MMP-2/TIMP-3. **(D)** MMP-2/TIMP-4. *\* P*-values were considered significant if p value (0.05) corrected for multiple comparisons by Bonferroni’s method (corrected p value=0.05/3=0.0166).

**Figure 5.**
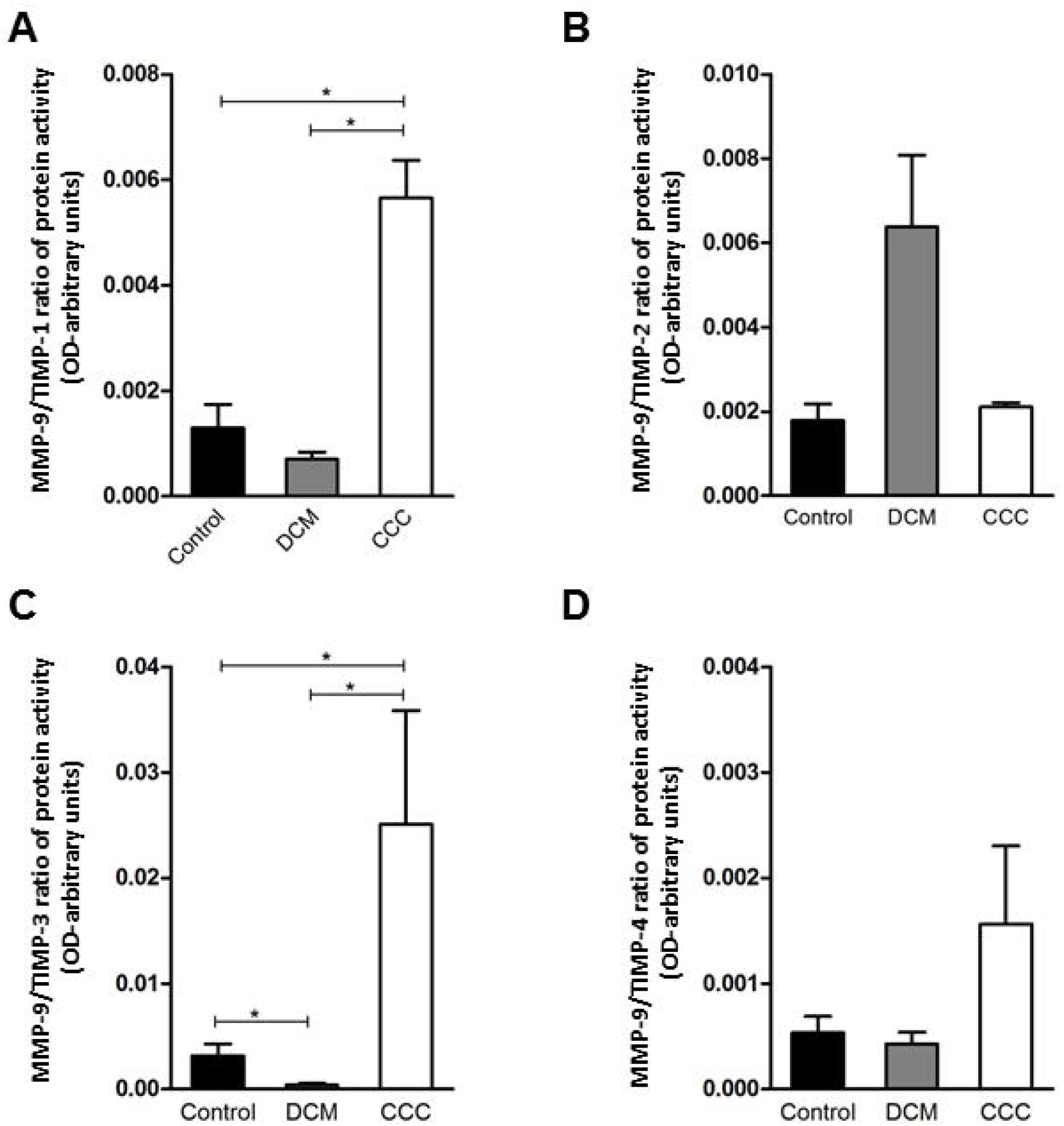
Ratios of MMP9/TIMP in heart tissue from CCC and DCM samples. Activity of MMP-9 / expression protein TIMP stoichiometric ratios were calculated for samples CCC, DCM and control **(A)** MMP-9/TIMP-1. **(B)** MMP-9/TIMP-2. **(C)** MMP-9/TIMP-3. **(D)** MMP-9/TIMP-4. Groups were compared by a non-parametrical test (Mann-Whitney Rank Sum Test) with GraphPad Prism software (version 6.0; GraphPad). Results were expressed as mean±SEM and arbitrary units. *\* P*-values were considered significant if p value (0.05) corrected for multiple comparisons by Bonferroni’s method (corrected p value=0.05/3=0.0166).

### Expression of MMP-3, MMP-8, MMP-12, MMP-13 and EMMPRIN

We evaluated gene and protein expression of the other classes of MMPs and EMMPRIN in myocardial samples from the CCC, DCM and control. **Figures 6A to 6C** shows that gene expression levels of MMP-12, MMP-13 and EMMPRIN were significantly higher in CCC and DCM myocardial tissue in comparison to samples from control group (P≤ 0.05). The gene expression of MMP-3 and MMP-8 were undetectable in all samples tested. There were no significant differences in protein expression of MMP-3, MMP-8, MMP-12, MMP-13 and EMMPRIN between CCC, DCM and control samples (**Figure 6D**).

**Figure 6.**
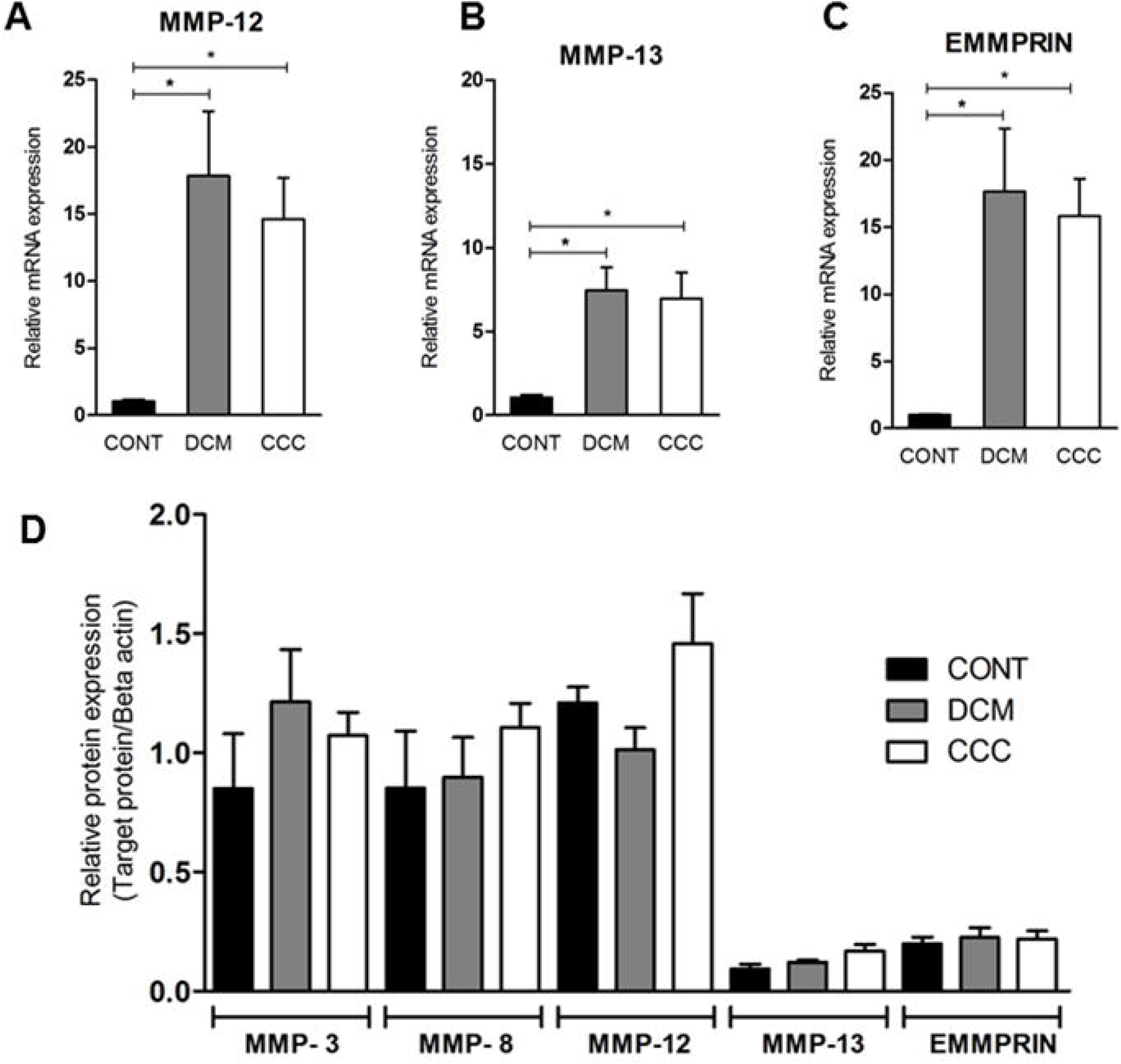
Expression of MMP-3, MMP-8, MMP-12, MMP-13 and EMMPRIN in heart tissue from CCC and DCM samples. Myocardial expression of MMP-12, MMP-13 and EMMPRIN mRNA. Real-time PCR analysis of mRNA expression in control (n=6), DCM (n=12) and CCC (n=17) myocardium. The expression was calculated as the mean ± SEM for each group as individual data points using the relative expression (fold change over control) by 2^−ΔΔCt^ method, normalized with the endogenous gene RPL0, as described in methods section. **(A)**Relative expression of MMP-mRNA. **(B)** Relative expression of MMP-13 mRNA. **(C)** Relative expression of EMMPRIN mRNA. The expression of mRNA of MMP-3 and MMP-8 was undetectable in all samples tested (data no show). **(D)** Densitometry analysis of the MMP-3, MMP-8, MMP-12, MMP-13 and EMMPRIN. Groups were compared by a non-parametrical test (Mann-Whitney Rank Sum Test) with GraphPad Prism software (version 6.0; GraphPad). Results were expressed as mean±SEM. *\* P*-values were considered significant if p value (0.05) corrected for multiple comparisons by Bonferroni’s method (corrected p value=0.05/3=0.0166).

## Discussion

Here, we report for the first time that the myocardium from CCC and DCM patients have alterations in MMP-2, MMP-9 and their TIMPs. We found that collagen area, MMP-2 and TIMP-2 protein levels and mRNA expression of the MMPs -2, -9, -12, -13, the TIMPs

-1, -2, -3, -4 and MMP inducer EMMPRIN were increased in both CCC and DCM samples as compared to controls. Importantly, MMP-9 protein levels and MMP-2 and MMP-9 enzymatic activity were increased only in CCC. Moreover, MMP Enzymatic activity/TIMP protein ratios MMP-2/TIMP-1, MMP-2/TIMP-3, MMP-9/TIMP-1 and MMP-9/TIMP-3 were increased in CCC as compared with DCM and control, and MMP-2/TIMP-2 and MMP-2/TIMP-4 ratios were higher among CCC than DCM.

Our findings on MMP-2 and MMP-9 corroborate the important involvement of MMPs in physiological and pathological myocardial remodeling [31–33]. Gutierrez et al. described an increase in MMP-2 and MMP-9 expression in hearts of mice acutely infected with *T. cruzi,* and TIMP treatment reduced myocarditis and increased survival [34]. Polyakova et al. demonstrated that increased expression of MMP-2 and MMP-9 is associated with collagen maturation in heart failure, showing an important role of these enzymes in fibrosis through collagen configuration, activation and deposition with cardiomyocyte hypertrophy and fibrosis contributing with remodeling and cardiac dysfunction [24]. Myocardial TNF-α was correlated with MMP-9 activity in a mouse model of myocardial infarction [35]. These authors demonstrated that TNF-α blockade decreased left ventricular dilation, which was associated with a decrease in the production and activity of MMP-9. Activation of TNF-α receptors can induce the synthesis of MMP-9 by activating the transcription factors, AP-1 (activator protein-1) and NF-kB (Nuclear factor kappa B), both capable of binding to MMP-9 promoter regions [36, 37]. Inflammatory cells present in the CCC myocardium produce high levels of TNF-α and other pro-inflammatory cytokines, like IFN-γ and IL-6 [38]. Human macrophages infected with *T. cruzi* produced increased levels of MMP-9, which was related to the production of cytokines such as IL-1β, TNF-α and IL-6 [39]. Medeiros et al. reported that neutrophils and monocytes from indeterminate and cardiac Chagas disease patients had higher intracellular levels of MMP-2 and MMP-9 than those found in non-infected individuals [40]. Together with our data showing increased protein levels of MMP-9 in CCC myocardium, this suggests that MMP-9 – possibly coming from inflammatory cells-might be the main MMP with activity induced by locally produced inflammatory cytokines. Our results that MMP-2 and MMP-9 activities are predominantly increased in CCC myocardium are consistent with the hypothesis that local inflammation induces metalloproteinase activity, exacerbating extracellular matrix remodeling with progressive fibrosis.

An imbalance in the proportions of MMPs and their endogenous TIMP inhibitors may lead to excessive degradation of ECM and disease [41]. The finding of increased ratios of MMP-2 and MMP-9 activity/TIMP protein levels suggest an overall dominance of MMP-2 and MMP-9 activity over TIMPs inhibition in myocardium samples from CCC but not DCM patients. The finding that only TIMP 1 and 3 showed significantly increased MMP/TIMP ratios in CCC vs both DCM and control might suggest TIMP-1 and TIMP-3 imbalance was key to the increased activity of both MMP-2 and -9 in CCC myocardium. Although all TIMPs have affinity for MMP-9, TIMP-1 was described to be the major endogenous inhibitor of MMP-9 [42]. Conversely, our data showing that TIMP-2 and TIMP-4 ratios were increased in CCC vs DCM in MMP-2 but not MMP-9 are consistent with TIMP-2 and TIMP-4 having an effect on MMP-2, but not on MMP-9. TIMP-2 is the most universal MMP inhibitor [24, 31], binding with high affinity to both the active and pro-form of MMP-2 [31]. Transcriptional regulation of MMP and TIMPs is partially overlapping; MMP-2 and TIMP-2 share the same transcription factor, AP-2 (activator protein) [31], which might explain the increased mRNA and protein expression of TIMP-2 and MMP-2 in myocardium samples from CCC and DCM patients.

Previous studies showed that patients with Chagas disease have higher circulating MMP-2 and MMP-9 protein and activity compared to patients with asymptomatic form of the disease [23, 43]. Sherbuck et al. showed that in patients with severe chagasic cardiomyopathy MMP-2 could be a predictive marker of mortality [44]. Fares et al. showed that CCC patients have a higher plasma levels and activity of MMP-2 and MMP-9 compared to patients with asymptomatic form of the disease, and they hypothesized that the increased activity of MMP-9 favors the development of the cardiac form of Chagas disease. Authors also noted that MMP-9 expression was positively correlated with the production of pro-inflammatory cytokines, like TNF-α and IL-1β [23]. It is likely that the increased tissue activities of MMP-2 and -9 are the proximal cause of the increased circulating levels of the same MMPs. Regarding the other MMP studied, the finding of increased mRNA of collagen I -degrading MMP-12, MMP-13 and MMP inducer EMMPRIN in CCC and DCM myocardial tissue in the absence of a corresponding increase in protein expression may be simply a response to similar transcriptional signals. Our study had limitations. The sample size was reduced and the ages and sex were not matched between the control and patient samples. This is linked to the fact that organ donors are almost always younger than the recipient cohort. However, in one of our previous studies [45], we had shown that these two covariates had a limited impact on the whole transcriptomic pattern.

Taken together, results suggest that myocardial remodeling and MMP-2 and MMP-9 activity are increased in CCC as compared with DCM, possibly by means of differential TIMP regulation of MMP activity. These findings may suggest a possible mechanism for the increased fibrosis, myocardial remodeling and cardiac dysfunction observed in patients with CCC as compared to other forms of cardiomyopathy. Pharmacological modulation of endogenous agents linked to fibrosis in CCC, like Galectin-3 [46] and microRNA-21 [47], or treatment with spironolactone [48], colchicine [49] or bone marrow cells [50] promoted a significant reduction in cardiac fibrosis in rodent models of Chagas disease, showing that pathological remodeling is a promising therapeutic target in CCC.

## Data Availability

All data produced in the present study are available upon reasonable request to the authors

## Acknowledgments

We thank all the patients and their relatives.

## Author contributions

Development of methodology and experimental work: MAB, LRPF, PCT, AISM, AFF, AK.

Histological analysis: RHBS, VD, LAB, FAG, FB, PP, FB.

Statistical analysis: MAB, PCT, ECN.

Writing, review, and/or revision of the manuscript: CC, JK, ECN.

## Supplemental materials

**Supplemental Figure 1.** Western blots showing MMP-3 (54 kD), -8 (65 kD), -12 (54 kD), -and EMMPRIN/CD147 (60 kD) protein bands. Protein bands stained with specific antibodies and developed as in Methods are depicted, together with the densitometric measurements using beta actin as a loading control.

**Supplemental Table 1.** Oligonucleotide primers used in gene expression analysis.

**Supplemental Table 2.** Histopathological features of the samples

