## Supplementary material for "Matrix metalloproteinase 2 and 9 enzymatic activities are selectively increased in the myocardium of Chronic Chagas Disease cardiomyopathy patients: role of TIMPs": suplemental materials: Supplemental Table_2.docx

**Supplemental Table 2.** Histopathological features of the samples

| **Etiology** | **Patient** | **Myocarditis ª** | **Hypertrophy ^b^** |
| --- | --- | --- | --- |
| CCC | 01 | 2/3+ | Y |
|  | 02 | 3+ | Y |
|  | 03 | 2/3+ | Y |
|  | 04 | 1+ | Y |
|  | 05 | 0/1+ | Y |
|  | 06 | 1+ | Y |
|  | 07 | 3+ | Y |
|  | 08 | 2/3+ | Y |
|  | 09 | 2+ | Y |
|  | 10 | 2+ | Y |
|  | 11 | 2/3+ | Y |
|  | 12 | 3+ | Y |
|  | 13 | 0 | Y |
|  | 14 | 3+ | Y |
|  | 15 | 3+ | Y |
|  | 16 | 2+ | Y |
|  | 17 | 3+ | Y |
| DCM | 18 | 0 | Y |
|  | 19 | 0 | Y |
|  | 20 | 0 | Y |
|  | 21 | 0 | Y |
|  | 22 | 0 | Y |
|  | 23 | 0 | Y |
|  | 24 | 1+ | Y |
|  | 25 | 0 | Y |
|  | 26 | 0 | Y |
|  | 27 | 0 | Y |
|  | 28 | 1+ | Y |
|  | 29 | 0 | Y |
| Control | 30 | 0 | N |
|  | 31 | 0 | N |
|  | 32 | 0 | N |
|  | 33 | 0 | N |
|  | 34 | 0 | N |
|  | 35 | 0 | N |

Y: Yes, N: No.

Control: Heart donors; DCM: Idiopathic dilated cardiomyopathy; CCC: Chronic Chagas cardiomyopathy.

ª Myocardites and Hypertrophy were rated by histopathology: (0: absent; 1+: mild; 2+: moderate, 3+: intense).

^b^ Cardiomyocyte hypertrophy was characterized by enlarged cells with prominent hyper chromatic nuclei.
