## Supplementary material for "Matrix metalloproteinase 2 and 9 enzymatic activities are selectively increased in the myocardium of Chronic Chagas Disease cardiomyopathy patients: role of TIMPs": suplemental materials: Supplemental_table_1.docx

**Supplemental table 1 :** Primers used in gene expression analysis.

| **Gene Name** | **Gene bank accession number** | **Sequence 5—3’** | **Amplicon (pb)** | **Concentration (nM)** |
| --- | --- | --- | --- | --- |
| MMP-2 | NM_001127891. 1 | CTGACCAAGGGTACAGCCTGTTAGGTGTAAATGGGTGCCATCAG | 108 | 200 |
| MMP-3 | NM_002422. 3 | AGACAGGCACTTTTGGCGCAAATGAAAACGAGGTCCTTGCTAG | 105 | 300 |
| MMP-8 | NM_002424. 2 | TGGAAACTCTGGACATGATGAAGAGATGCTCATTTTGATGCCG | 117 | 300 |
| MMP-9 | NM_004994. 2 | GATGGGAAGTACTGGCGATTCTCCCAGTTTGCAGAGCGCTACC | 116 | 200 |
| MMP-12 | NM_002426.4 | TACGCAATCCGGAAAGCTTTCATGAGCTCCACGGGCAAAA | 111 | 200 |
| MMP-13 | NM_002427. 3 | GGAATTAAGGAGCATGGCGACCCATAATTTGGCCCAGGAGG | 115 | 300 |
| EMMPRIN | NM_001728. 3 | CTCACCTGCTCCTTGAATGACAGCACCTTGAACTCCGTTTTCTGG | 114 | 300 |
| TIMP-1 | NM_003254. 2 | GCTGTGAGGAATGCACAGTGTTTCCTTTTCAGAGCCTTGGAGGA | 105 | 200 |
| TIMP-2 | NM_003255. 4 | AGTGCAAGATCACGCGCTGTGGTGCCCGTTGATGTTCTT | 103 | 200 |
| TIMP-3 | NM_000362. 4 | CTGACAGGTCGCGTCTATGATGAGGTGATACCGATAGTTCAGCCC | 113 | 200 |
| TIMP-4 | NM_003256. 3 | CTGCCAAATCACCACCTGCTACATAGAGCTTTCGTTCCAACAGC | 95 | 200 |
| RECK | NM_021111. 2. | CTCTGCAGATTGAAGCCTGCACCTGGGAAATGATGAGGGC | 114 | 200 |
| RPLP0 | NM_Q3B7A4 | GAAATCCTGAGTGATGATGTGCAGGATGACGAGCGGAAAGGAGAA | 103 | 200 |
