## Supplementary figures and images for "Matrix metalloproteinase 2 and 9 enzymatic activities are selectively increased in the myocardium of Chronic Chagas Disease cardiomyopathy patients: role of TIMPs"

### Supplemental_figure_1.TIF

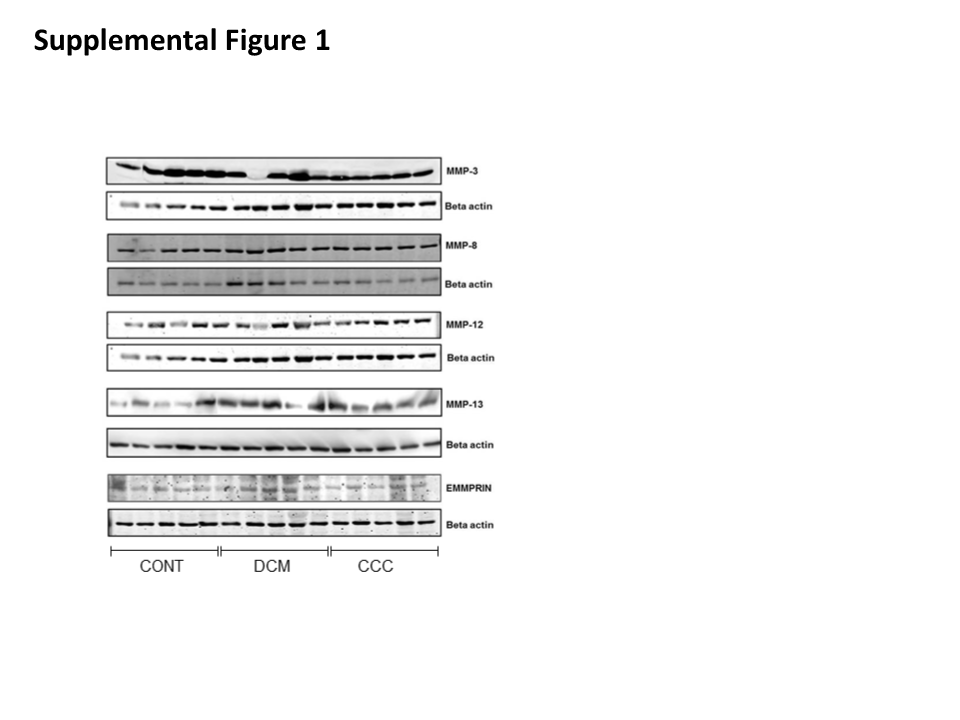
